# Moving beyond overall cesarean rates: Evaluating associations of delivery mode and adverse outcomes by Robson Group among the PRISMA Maternal and Newborn Health Study cohort in sub-Saharan Africa and South Asia

**DOI:** 10.64898/2026.08.18.26360699

**Authors:** Sasha G. Baumann, Nida S. Yazdani, A James, Vanessa Amabo, Rupa Talukdar, Blair J. Wylie, Victor Akelo, Florence Aweyo, Santosh Joseph Benjamin, Anne George Cherian, Zahra Hoodbhoy, Margaret P. Kasaro, Poonam Kataria, Sarmila Mazumder, Christopher N. Mores, Wilbroad Mutale, Muhammad Imran Nisar, Kaweeta Kumari, Bushra Liaqat, Erin M. Oakley, Caleb Sagam, Neeraj Sharma, Emily R. Smith, Nazia Binte Ali, M. Bridget Spelke

**Affiliations:** Department of Global Health, George Washington University, Washington, D.C., United States; Department of Paediatrics and Child Health, The Aga Khan University, Karachi, Pakistan; Community Medicine, Christian Medical College Vellore, Vellore, India; East Carolina Brody School of Medicine, Greenville, N.C., United States; Implementation Science Domain, Society for Applied Studies, New Delhi, India; Department of Obstetrics and Gynecology, Beth Israel Deaconess Medical Center, Boston, M.A., United States; Center for Global Health Research, Kenya Medical Research Institute, Kisumu, Kenya; Liverpool School of Tropical Medicine, Liverpool, England; Obstetrics and Gynecology, Christian Medical College Vellore, Vellore, India; UNC Global Projects Zambia, Lusaka, Zambia; Department of Health Policy and Management, University of Zambia School of Public Health, Lusaka, Zambia; Department of Obstetrics and Gynecology, Creek General Hospital, Karachi, Pakistan; Department of Obstetrics and Gynecology, Koohi Goth Hospital, Karachi, Pakistan; Department of Obstetrics and Gynecology, University of North Carolina at Chapel Hill, Chapel Hill, N.C., United States

**Keywords:** Robson Groups, cesarean section, birth outcomes, delivery mode, LMICs, adverse outcomes, maternal outcomes, Robson Classification

## Abstract

**Objective:** Rising cesarean section (CS) rates in low- and middle-income countries may mask a triple burden of unmet need, overuse, and unsafe provision. Using the Robson Ten-Group Classification, a global standard for monitoring and comparing institutional deliveries, we examine CS incidence and associations with adverse outcomes.

**Methods:** Data were drawn from the Pregnancy Risk, Infant Surveillance, and Measurement Alliance Maternal and Newborn Health Study, an open cohort study conducted from 2022 to 2025 in Kenya, Zambia, India, and Pakistan. We generated descriptive statistics for Robson Groups and within-group relative risks of adverse events for CS versus vaginal delivery using multivariable adjusted log Poisson models.

**Results:** Among 10,996 women, 29% delivered by CS. Group 5 (prior CS) and Group 10 (preterm) were the largest contributors to CS, accounting for 28% and 17% of all CS deliveries, respectively. Between-site differences in CS incidence were most pronounced for Groups 2 and 4 (induced labor/pre-labor CS), ranging from 20-60% for nullipara and 7-44% for multipara. Compared to vaginal delivery, CS was associated with increased risk of maternal near-miss, prolonged hospitalization, hemorrhage, and newborn intensive care unit admission. These associations differed in magnitude when stratified by Robson Group, with the greatest risk among lower-risk groups.

**Conclusion:** Repeat CS, preterm deliveries, labor induction, and pre-labor CS were key drivers of CS, with considerable differences between sites. Equipping facilities to safely manage labor induction, trials of labor after cesarean, and preterm deliveries is critical to improving quality of care.

## INTRODUCTION

### Background

Global cesarean section (CS) rates rose from 7% in 1990 to 21% in 2021, with projections showing a continued rise of 4% annually [1, 2]. In many low- and middle-income countries (LMICs), unmet need for CS persists alongside non-medically indicated or poor quality procedures, imposing a triple burden of underuse, overuse, and unsafe provision that poses risks to patients and strains health systems [3, 4]. Interpreting CS trends is further complicated by the growing prevalence of non-communicable diseases in pregnancy, such as hypertension and diabetes, which contribute to the likelihood of surgical delivery [5, 6].

Though population-level CS rates are used for benchmarking, the World Health Organization (WHO) does not recommend a single target CS rate. Ecological analyses have suggested that population-level CS rates below 9% may reflect underuse, while rates above 15-16% are not associated with reductions in maternal or neonatal mortality after adjustment for socioeconomic factors [7–9]. These thresholds should be interpreted cautiously, as they do not account for differences in case-mix complexity, referral patterns, or maternal risk profiles.

The Robson Ten-Group Classification System provides a standardized method for categorizing deliveries into mutually exclusive groups based on parity, prior CS, labor onset, fetal number, gestational age, and fetal presentation (**Figure 1**) [10]. By stratifying CS rates according to clinically meaningful criteria, the Robson Classification enables the evaluation of practice patterns and trends across facilities and over time [11]. The Robson Classification has been largely utilized in high-income settings, in part due to its reliance on accurate gestational age dating, which remains a challenge in LMICs [4].

**Figure 1.**
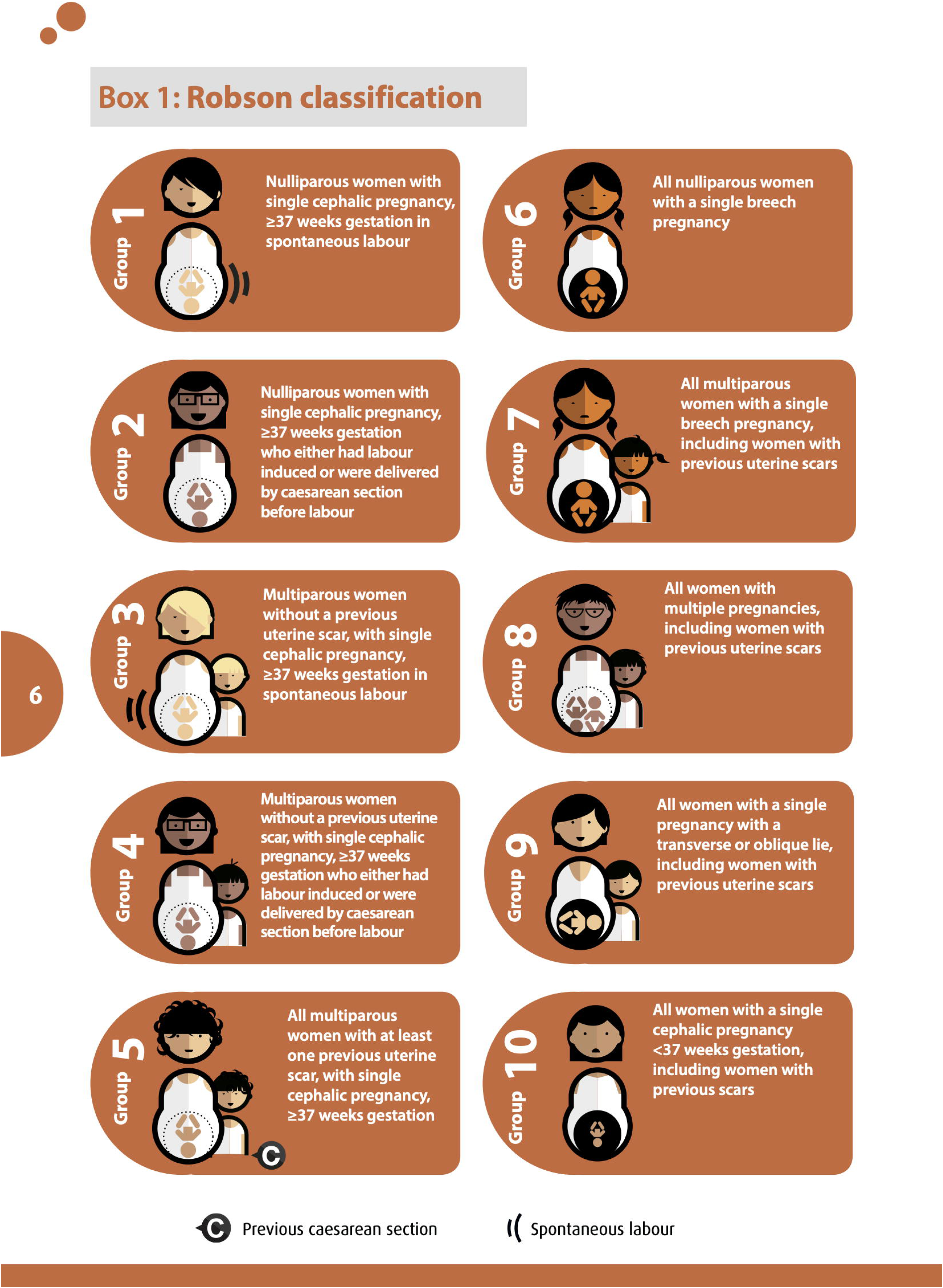
Robson 10-Group Classification System. Image reproduced from the World Health Organization Statement on Caesarean Section Rates WHO/RHR/15.02. 2015.

Previous research demonstrates significant differences in CS rates by Robson Classification group [12]. Across diverse settings, Group 5 (prior CS) consistently contributes the largest share of CS, accounting for 15-50% of CS deliveries and reflecting the domino effect from primary to repeat CS [13]. Groups 1 and 2 (nullipara with spontaneous labor or induced labor/pre-labor CS, respectively) account for 30-50% of CS [14]. Most studies using the Robson Classification focus on group size and contribution to overall CS rate; data linking delivery mode to maternal and neonatal morbidity and mortality within Robson Groups remain sparse. Without these data, interpretation of whether observed CS patterns in LMICs reflect appropriate, unsafe, or insufficient use is limited [15].

### Rationale and Objectives

Maternal risk profiles are shifting across LMICs, with hypertensive disorders, diabetes, anemia, and obesity shaping demand for surgical delivery, necessitating risk-stratified evaluation to determine appropriate CS use within comparable populations [16, 17]. We calculate CS incidence and relative contribution by Robson Group in Kenya, Zambia, India, and Pakistan. We further explore association between delivery mode and adverse maternal and infant outcomes by Robson Group, offering new insights into obstetric practice in an underrepresented context.

## MATERIALS AND METHODS

### Study design and population

We use data from the Pregnancy Risk, Infant Surveillance, and Measurement Alliance (PRISMA) Maternal and Newborn Health Study: a prospective cohort study in Kenya, Zambia, Ghana, India, and Pakistan. Study procedures have been described previously [18]. Briefly, study assessments were conducted at <20, 20, 28, 32, and 36 weeks of gestation, at delivery or within three days postpartum, and at one, four, and six weeks, six months, and one year postpartum. Labor and delivery information was collected by trained staff through observation, maternal recall, and/or record abstraction. This analysis included participants with pregnancy duration ≥28 weeks, birthweight ≥1000 grams, institutional delivery, confirmed mode of delivery, and who completed a six-week postpartum follow-up visit. We restricted analysis to participants enrolled between 2022 and 2025 and excluded Ghana site data due to missing data at time of analysis. Enrollment began in September 2022 in Pakistan, November 2022 in Kenya, December 2022 in Zambia, July 2023 in South India, and December 2023 in North India.

### Variable definitions

Robson Groups were defined according to the WHO Implementation Manual, based on parity, previous CS, fetal presentation, number of fetuses, gestational age, and labor onset [11]. All participants completed an ultrasound at <20 weeks gestation. Gestational age at delivery was determined by best obstetric estimate, which combines early-pregnancy ultrasound dating and reported date of last menstrual period. Parity and previous CS were self-reported. Labor onset was reported as spontaneous, induced, or pre-labor CS.

We included adverse outcomes that occurred between delivery and six weeks postpartum (case definitions in **Table S1**). Maternal outcomes included prolonged hospitalization, severe postpartum hemorrhage, maternal near-miss, and postpartum depression. Infant adverse outcomes included neonatal death, possible severe bacterial infection, and neonatal intensive care unit (NICU) admission.

### Statistical analysis

Robson group sizes, CS incidence, and the absolute and relative contributions of each group to the overall proportion of CS were calculated for all sites combined and separately, in line with the Robson Classification Implementation Manual [11]. We calculated within-group associations of CS with adverse outcomes using Generalized Linear Models with log link and Poisson distribution and robust standard error, adjusting for maternal age group, educational attainment category, body mass index at enrollment, height, wealth quintile, and study site. Statistical analyses were performed using Stata 18.5 MP-Parallel Edition and R version 4.2.2 software.

### Ethics approval and consent

Approvals were received from the following institutional review boards (IRBs) and ethical review committees (ERCs): India (Office of Research, Christian Medical College, Vellore: IRB14553; ERC, Society for Applied Studies, New Delhi: SAS/ERC/ReMAPPStudy/2022; Government of India, Department of Health Research: EC/NEW/INST/2022/DL/0140); Kenya (KEMRI Scientific and Ethics Review Unit: KEMRI/RES/7/3/1; Liverpool School of Tropical Medicine Ethics Committee: RGETEM044; County Government of Kisumu Department of Health: IERC.1B/VOL.II/549/21; Republic of Kenya National Commission for Science, Technology, and Innovation: NACOSTI-P-22-16990); Pakistan (The Aga Khan University ERC: 2022-5920- 22763; Pakistan National Institutes of Health, National Bioethics Committee: 4-87/NBC- 588/22/187NBCR); Zambia (University of North Carolina Chapel Hill Office of Human Research Ethics: 356795; University of Zambia Biomedical Research Ethics Committee: 016-04-14); and the United States (George Washington University IRB: NCR224396; Columbia University IRB: IRB-AAAU7504; Harvard University IRB: IRB23-1093; University of Alabama at Birmingham IRB: IRB-300013081). Informed consent or assent with parental consent was requested.

## RESULTS

We analyzed data for 2,242 women in Kenya, 1,415 in Zambia, 1,828 in North India, 1,919 in South India, and 3,592 in Pakistan (N=10,996; **Figure 2**). **Table 1** presents a descriptive snapshot of facility-level delivery patterns for one month in 2025 (intended as contextual information). Most women were 20-34 years (84%), completed secondary education or higher (72%), and were multiparous (68%) (**Table 2, Table S2**). Thirteen percent reported a previous cesarean. Among current pregnancies, 97% were singleton, 96% cephalic, and 85% delivered at term. Provider-initiated delivery varied by site: labor induction prevalence ranged from 17% in Kenya to 47% in South India, and pre-labor CS from 3% in Kenya to 19% in Pakistan. CS incidence was 20% Kenya, 26% in North and South India, 33% in Zambia, and 36% in Pakistan.

**Figure 2.**
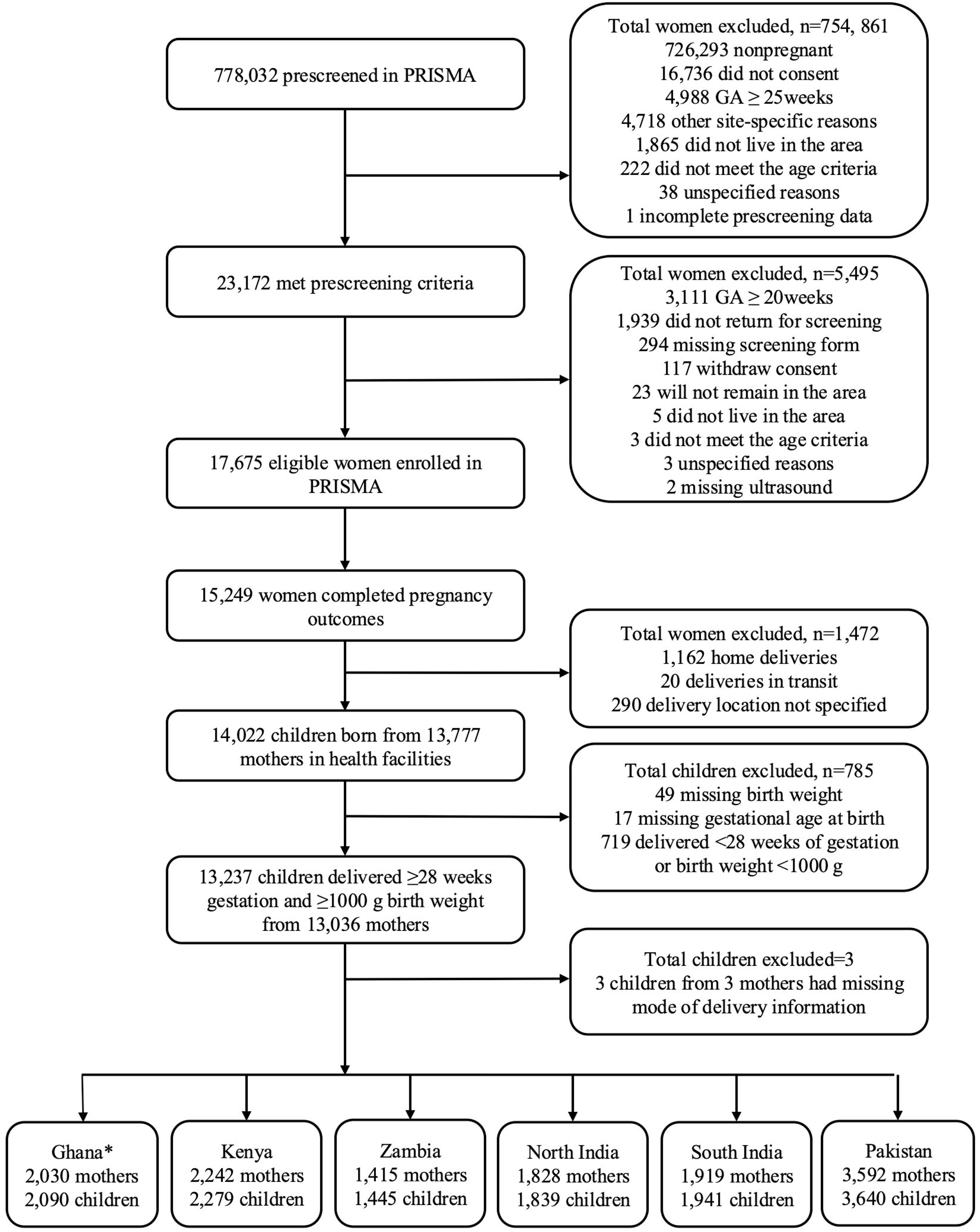
Flowchart of analytical sample selection (*Ghana data excluded from analysis due to lack of information on previous cesarean section)

**Table 1.** Snapshot of primary study facility level, staffing, and cesarean section rates.

| Site | Facility Information |  |  | Staffing (FT=full time; PT=part time) |  |  | PRISMA Deliveries (%)<sup>2</sup> |
| --- | --- | --- | --- | --- | --- | --- | --- |
|  | Level | Ownership | CS Rate (%)<sup>1</sup> | Obstetricians | Midwives | Surgeons |  |
| Kenya | Tertiary | Government | 36% | 8 | 19 | 16 | 20% |
|  | Secondary | Government | 35% | 3 | 15 | 3 | 10% |
|  | Secondary | Government | 10% | 1 |  | 0 | 21% |
|  | Secondary | Government | 36% | 2 | 6 FT, 8 PT | 0 | 22% |
| Pakistan | Secondary | Charitable | 39% | 15 | 31 | 10 | 36% |
|  | Tertiary | Private | 33% | 32 | 20 | 18 | 14% |
| South India | Secondary | Private | 17% | 3 | 58 | 1 PT | 75% |
|  | Tertiary | Private | 41% | 18 | 75 | 18 | 10% |
| North India | Secondary | Government | 7% | 2 | 23 | 0 | 31% |
|  | Secondary | Government | 30% | 3 | 35 | 1 | 11% |
|  | Secondary | Private | 50% | 1 FT, 1 PT | 6 | 1 PT | 16% |
| Zambia | Tertiary | Government | 53% | 29 FT, 0 PT | 354 FT, 44 PT | No data | 80% |
|  | Primary | Government | 18% | 2 FT, 0 PT | 189 FT, 46 PT | 0 | 20% |

<sup>1</sup>Facility-level data reflect one month of routine service statistics from September, October, or November 2025. They do not correspond to the full period of participant deliveries in the cohort.
<sup>2</sup>Percentage of all PRISMA cohort participant deliveries that occurred at the facility. Facilities with few PRISMA deliveries not shown.

**Table 2.**
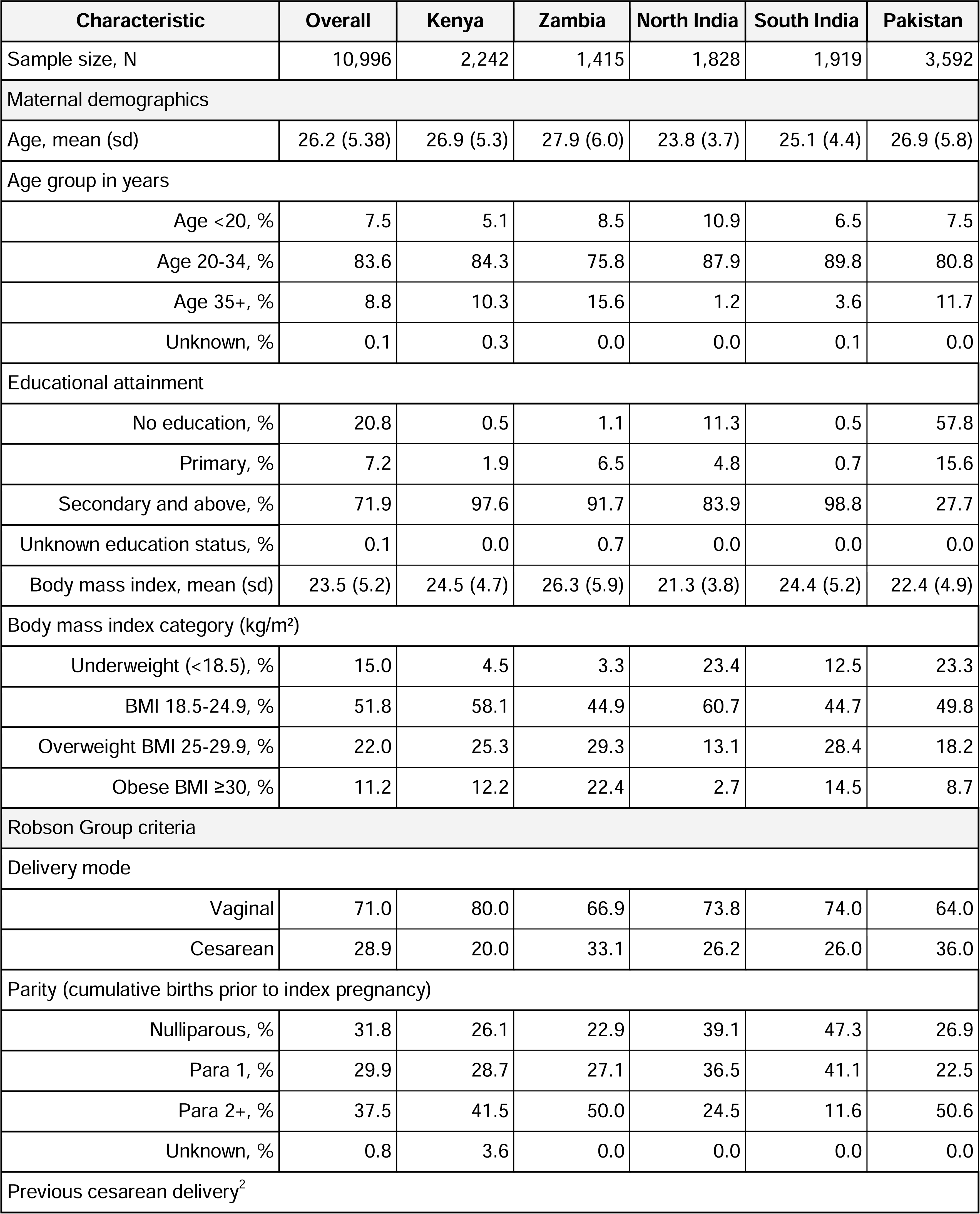
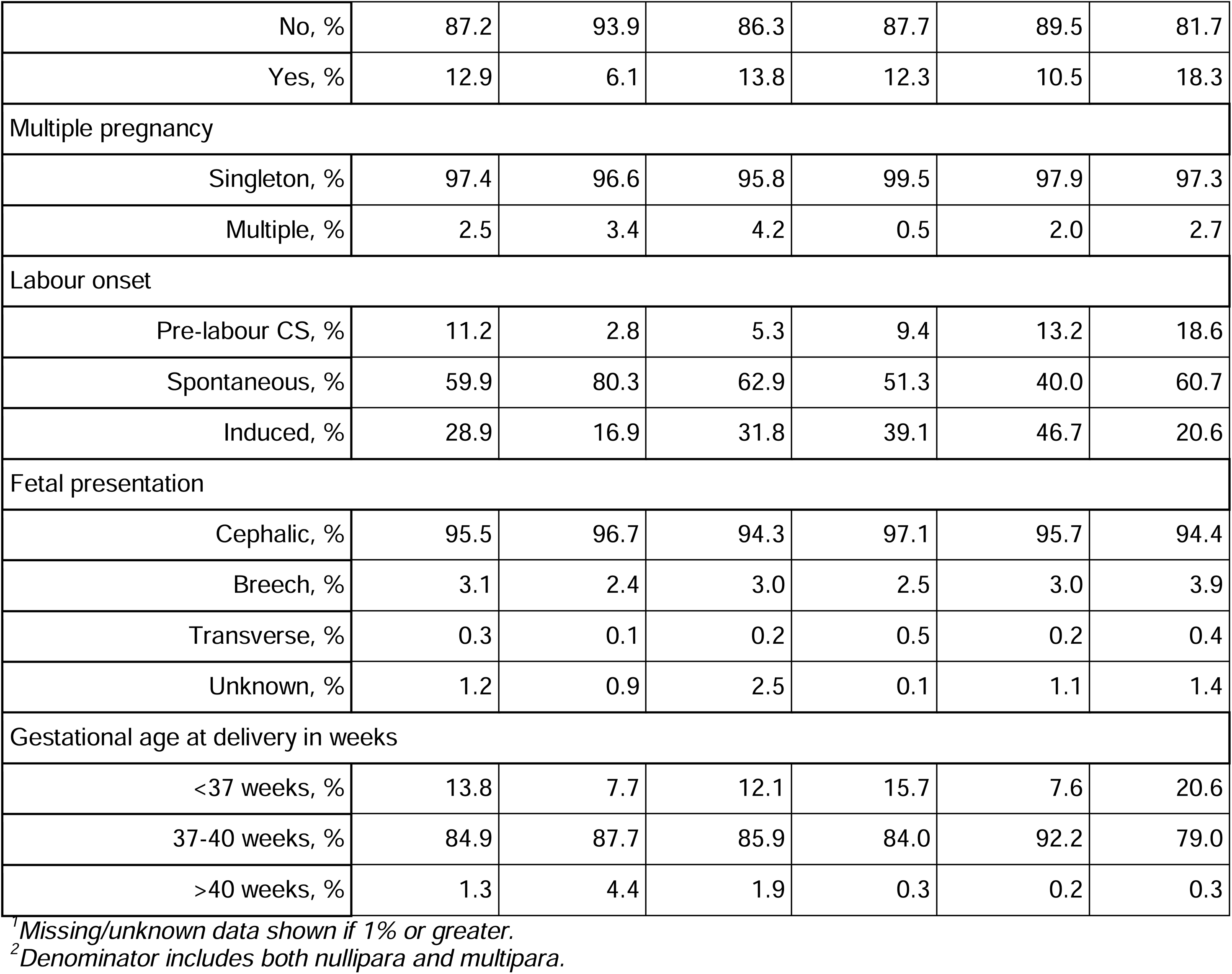
Characteristics of analytical sample.

| Characteristic | Overall | Kenya | Zambia | North India | South India | Pakistan |
| --- | --- | --- | --- | --- | --- | --- |
| Sample size, N | 10,996 | 2,242 | 1,415 | 1,828 | 1,919 | 3,592 |
| Maternal demographics |  |  |  |  |  |  |
| Age, mean (sd) | 26.2 (5.38) | 26.9 (5.3) | 27.9 (6.0) | 23.8 (3.7) | 25.1 (4.4) | 26.9 (5.8) |
| Age group in years |  |  |  |  |  |  |
| Age <20, % | 7.5 | 5.1 | 8.5 | 10.9 | 6.5 | 7.5 |
| Age 20-34, % | 83.6 | 84.3 | 75.8 | 87.9 | 89.8 | 80.8 |
| Age 35+, % | 8.8 | 10.3 | 15.6 | 1.2 | 3.6 | 11.7 |
| Unknown, % | 0.1 | 0.3 | 0.0 | 0.0 | 0.1 | 0.0 |
| Educational attainment |  |  |  |  |  |  |
| No education, % | 20.8 | 0.5 | 1.1 | 11.3 | 0.5 | 57.8 |
| Primary, % | 7.2 | 1.9 | 6.5 | 4.8 | 0.7 | 15.6 |
| Secondary and above, % | 71.9 | 97.6 | 91.7 | 83.9 | 98.8 | 27.7 |
| Unknown education status, % | 0.1 | 0.0 | 0.7 | 0.0 | 0.0 | 0.0 |
| Body mass index, mean (sd) | 23.5 (5.2) | 24.5 (4.7) | 26.3 (5.9) | 21.3 (3.8) | 24.4 (5.2) | 22.4 (4.9) |
| Body mass index category (kg/m<sup>2</sup>) |  |  |  |  |  |  |
| Underweight (<18.5), % | 15.0 | 4.5 | 3.3 | 23.4 | 12.5 | 23.3 |
| BMI 18.5-24.9, % | 51.8 | 58.1 | 44.9 | 60.7 | 44.7 | 49.8 |
| Overweight BMI 25-29.9, % | 22.0 | 25.3 | 29.3 | 13.1 | 28.4 | 18.2 |
| Obese BMI ≥30, % | 11.2 | 12.2 | 22.4 | 2.7 | 14.5 | 8.7 |
| Robson Group criteria |  |  |  |  |  |  |
| Delivery mode |  |  |  |  |  |  |
| Vaginal | 71.0 | 80.0 | 66.9 | 73.8 | 74.0 | 64.0 |
| Cesarean | 28.9 | 20.0 | 33.1 | 26.2 | 26.0 | 36.0 |
| Parity (cumulative births prior to index pregnancy) |  |  |  |  |  |  |
| Nulliparous, % | 31.8 | 26.1 | 22.9 | 39.1 | 47.3 | 26.9 |
| Para 1, % | 29.9 | 28.7 | 27.1 | 36.5 | 41.1 | 22.5 |
| Para 2+, % | 37.5 | 41.5 | 50.0 | 24.5 | 11.6 | 50.6 |
| Unknown, % | 0.8 | 3.6 | 0.0 | 0.0 | 0.0 | 0.0 |
| Previous cesarean delivery<sup>2</sup> |  |  |  |  |  |  |
| No, % | 87.2 | 93.9 | 86.3 | 87.7 | 89.5 | 81.7 |
| Yes, % | 12.9 | 6.1 | 13.8 | 12.3 | 10.5 | 18.3 |
| Multiple pregnancy |  |  |  |  |  |  |
| Singleton, % | 97.4 | 96.6 | 95.8 | 99.5 | 97.9 | 97.3 |
| Multiple, % | 2.5 | 3.4 | 4.2 | 0.5 | 2.0 | 2.7 |
| Labour onset |  |  |  |  |  |  |
| Pre-labour CS, % | 11.2 | 2.8 | 5.3 | 9.4 | 13.2 | 18.6 |
| Spontaneous, % | 59.9 | 80.3 | 62.9 | 51.3 | 40.0 | 60.7 |
| Induced, % | 28.9 | 16.9 | 31.8 | 39.1 | 46.7 | 20.6 |
| Fetal presentation |  |  |  |  |  |  |
| Cephalic, % | 95.5 | 96.7 | 94.3 | 97.1 | 95.7 | 94.4 |
| Breech, % | 3.1 | 2.4 | 3.0 | 2.5 | 3.0 | 3.9 |
| Transverse, % | 0.3 | 0.1 | 0.2 | 0.5 | 0.2 | 0.4 |
| Unknown, % | 1.2 | 0.9 | 2.5 | 0.1 | 1.1 | 1.4 |
| Gestational age at delivery in weeks |  |  |  |  |  |  |
| <37 weeks, % | 13.8 | 7.7 | 12.1 | 15.7 | 7.6 | 20.6 |
| 37-40 weeks, % | 84.9 | 87.7 | 85.9 | 84.0 | 92.2 | 79.0 |
| >40 weeks, % | 1.3 | 4.4 | 1.9 | 0.3 | 0.2 | 0.3 |

<sup>1</sup>Missing/unknown data shown if 1% or greater.
<sup>2</sup>Denominator includes both nullipara and multipara.

Robson Group sizes for the sample as a whole are presented in **Table 3** and for each site in **Table S3**. Throughout the results, each Robson Group is referenced by its most salient characteristic; definitions are provided in **Figure 1**. Overall, 32% of women were classified as Group 3 (multipara with spontaneous labor), 16% as Group 1 (nullipara with spontaneous labor), 15% as Group 4 (multipara with induced labor or prelabor CS), and 12% as Group 10 (preterm deliveries). Combined, Groups 3 and 4 accounted for 46% of the sample. Relative group size differed between sites, with the greatest difference for Group 3, which constituted 49% of the sample in Kenya versus 19% in South India. This was followed by Group 2 size variability, ranging from 4% in Kenya to 27% in South India.

**Table 3.**
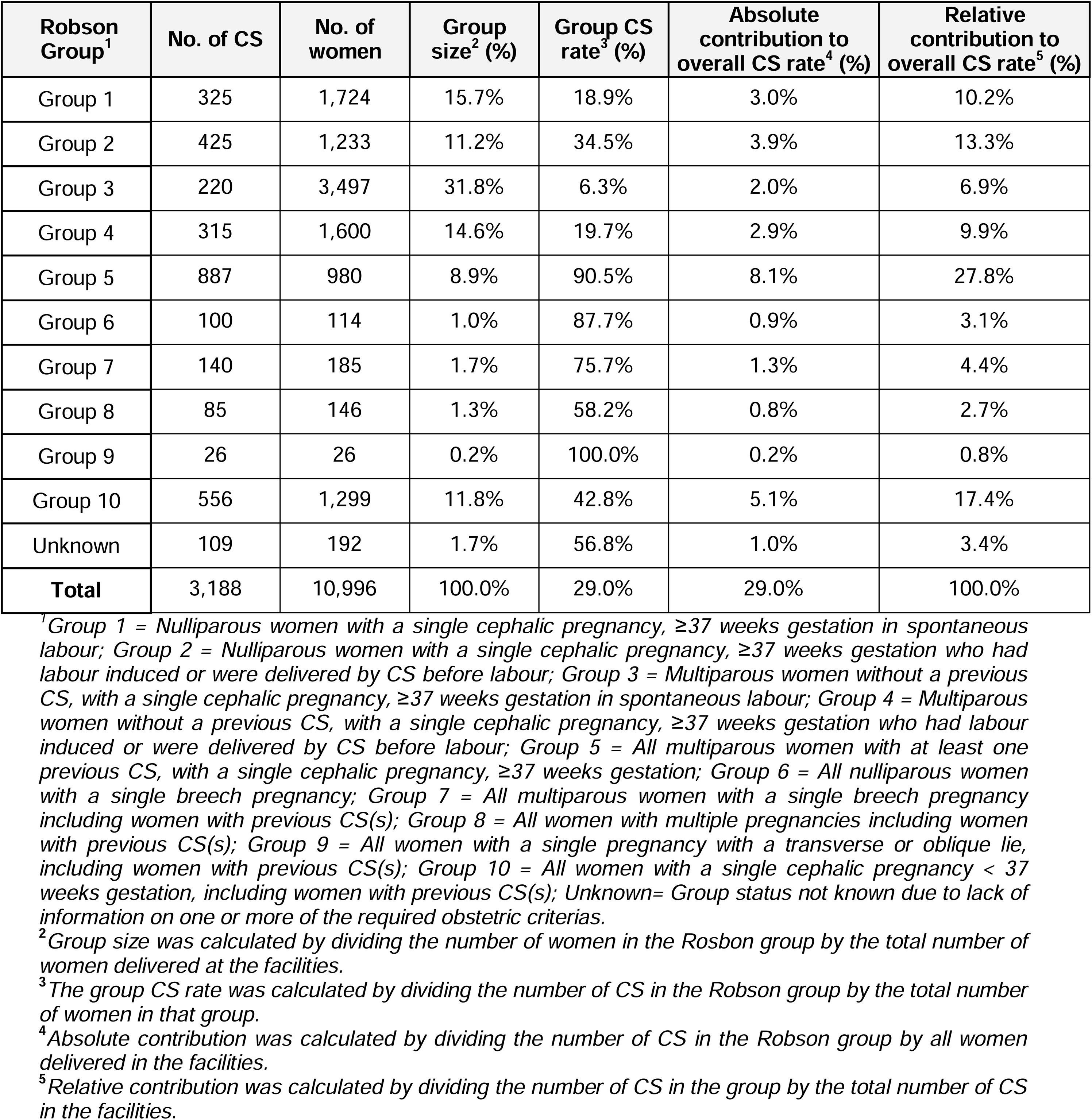
Robson Group size and cesarean section (CS) rates for all sites combined.

| Robson Group<sup>1</sup> | No. of CS | No. of women | Group size<sup>2</sup> (%) | Group CS rate<sup>3</sup> (%) | Absolute contribution to overall CS rate<sup>4</sup> (%) | Relative contribution to overall CS rate<sup>5</sup> (%) |
| --- | --- | --- | --- | --- | --- | --- |
| Group 1 | 325 | 1,724 | 15.7% | 18.9% | 3.0% | 10.2% |
| Group 2 | 425 | 1,233 | 11.2% | 34.5% | 3.9% | 13.3% |
| Group 3 | 220 | 3,497 | 31.8% | 6.3% | 2.0% | 6.9% |
| Group 4 | 315 | 1,600 | 14.6% | 19.7% | 2.9% | 9.9% |
| Group 5 | 887 | 980 | 8.9% | 90.5% | 8.1% | 27.8% |
| Group 6 | 100 | 114 | 1.0% | 87.7% | 0.9% | 3.1% |
| Group 7 | 140 | 185 | 1.7% | 75.7% | 1.3% | 4.4% |
| Group 8 | 85 | 146 | 1.3% | 58.2% | 0.8% | 2.7% |
| Group 9 | 26 | 26 | 0.2% | 100.0% | 0.2% | 0.8% |
| Group 10 | 556 | 1,299 | 11.8% | 42.8% | 5.1% | 17.4% |
| Unknown | 109 | 192 | 1.7% | 56.8% | 1.0% | 3.4% |
| **Total** | **3,188** | **10,996** | **100.0%** | **29.0%** | **29.0%** | **100.0%** |

<sup>1</sup>Group 1 = Nulliparous women with a single cephalic pregnancy, ≥37 weeks gestation in spontaneous labour; Group 2 = Nulliparous women with a single cephalic pregnancy, ≥37 weeks gestation who had labour induced or were delivered by CS before labour; Group 3 = Multiparous women without a previous CS, with a single cephalic pregnancy, ≥37 weeks gestation in spontaneous labour; Group 4 = Multiparous women without a previous CS, with a single cephalic pregnancy, ≥37 weeks gestation who had labour induced or were delivered by CS before labour; Group 5 = All multiparous women with at least one previous CS, with a single cephalic pregnancy, ≥37 weeks gestation; Group 6 = All nulliparous women with a single breech pregnancy; Group 7 = All multiparous women with a single breech pregnancy including women with previous CS(s); Group 8 = All women with multiple pregnancies including women with previous CS(s); Group 9 = All women with a single pregnancy with a transverse or oblique lie, including women with previous CS(s); Group 10 = All women with a single cephalic pregnancy < 37 weeks gestation, including women with previous CS(s); Unknown= Group status not known due to lack of information on one or more of the required obstetric criterias.
<sup>2</sup>Group size was calculated by dividing the number of women in the Robson group by the total number of women delivered at the facilities.
<sup>3</sup>The group CS rate was calculated by dividing the number of CS in the Robson group by the total number of women in that group.
<sup>4</sup>Absolute contribution was calculated by dividing the number of CS in the Robson group by all women delivered in the facilities.
<sup>5</sup>Relative contribution was calculated by dividing the number of CS in the group by the total number of CS in the facilities.

We calculated CS incidence by Robson Group overall and for each site (**Table 3**, **Figure 3, Table S4).** Group 6 (nullipara with breech pregnancy) had the greatest between-site differences in CS incidence, with the lowest in Kenya (50%) and the other sites falling between 74-100%. This was followed by Group 2 (nullipara with induced labour or pre-labor CS), within which both Indian sites had the lowest CS incidences at 20% and 28%, compared with 46-60% for Pakistan, Zambia, and Kenya. Similarly, for Group 4 (multipara with induced labor or pre-labor CS), Zambia and Kenya reported the highest CS incidence at 44% and 34%, compared to 7% and 10% in North and South India. Relative contributions of each Robson Group to overall CS incidence are visualized in **Figure 4**. Group 5 (prior CS) was the largest contributor to CS at all sites, followed by Group 4 (multipara with induced labor onset or prelabor CS) in Zambia, Kenya, and South India, and Group 10 (preterm deliveries) in Pakistan and North India.

**Figure 3.**
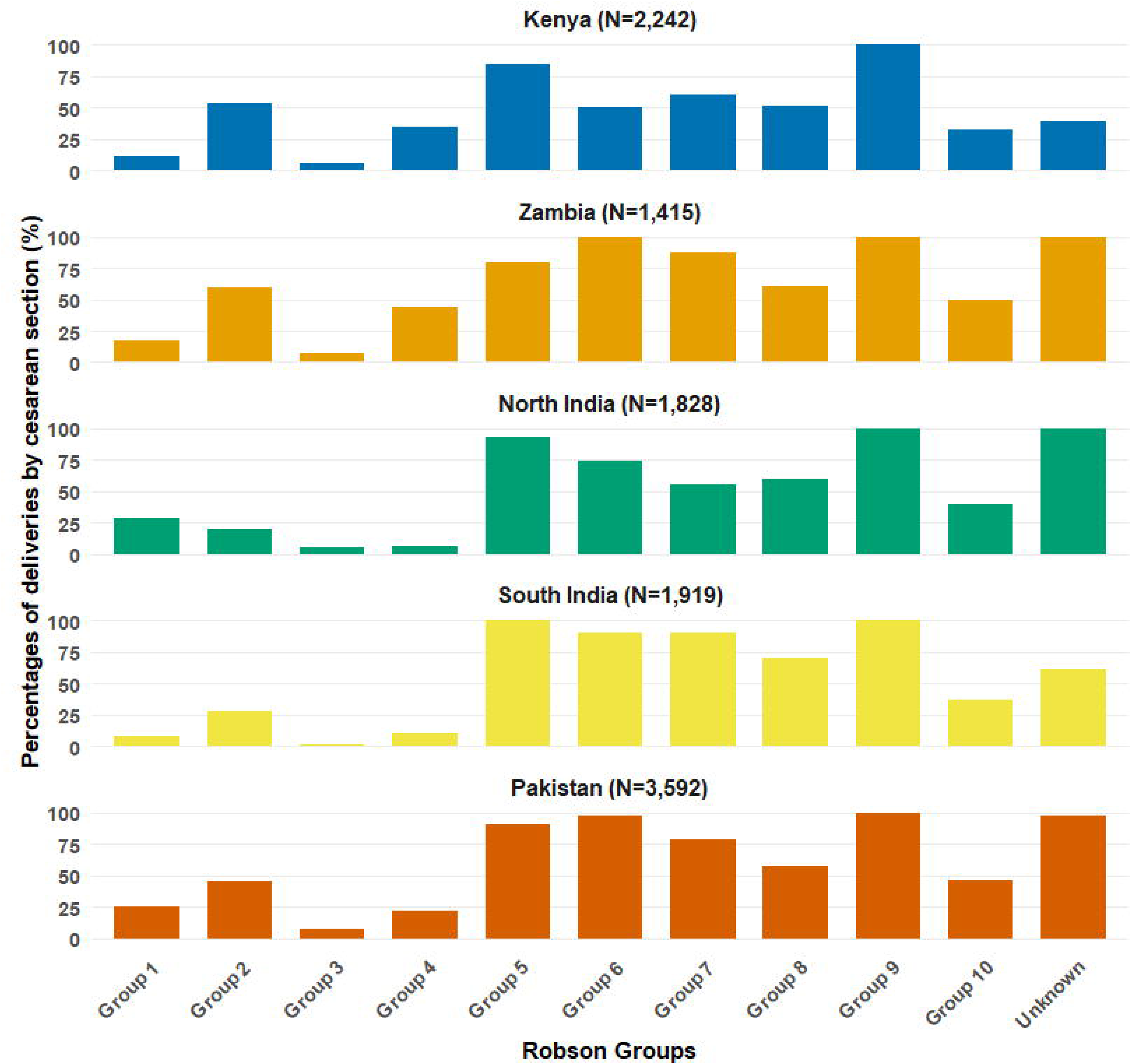
Proportion of deliveries by cesarean section within each Robson Group, stratified by study site (N=10,996)

**Figure 4.**
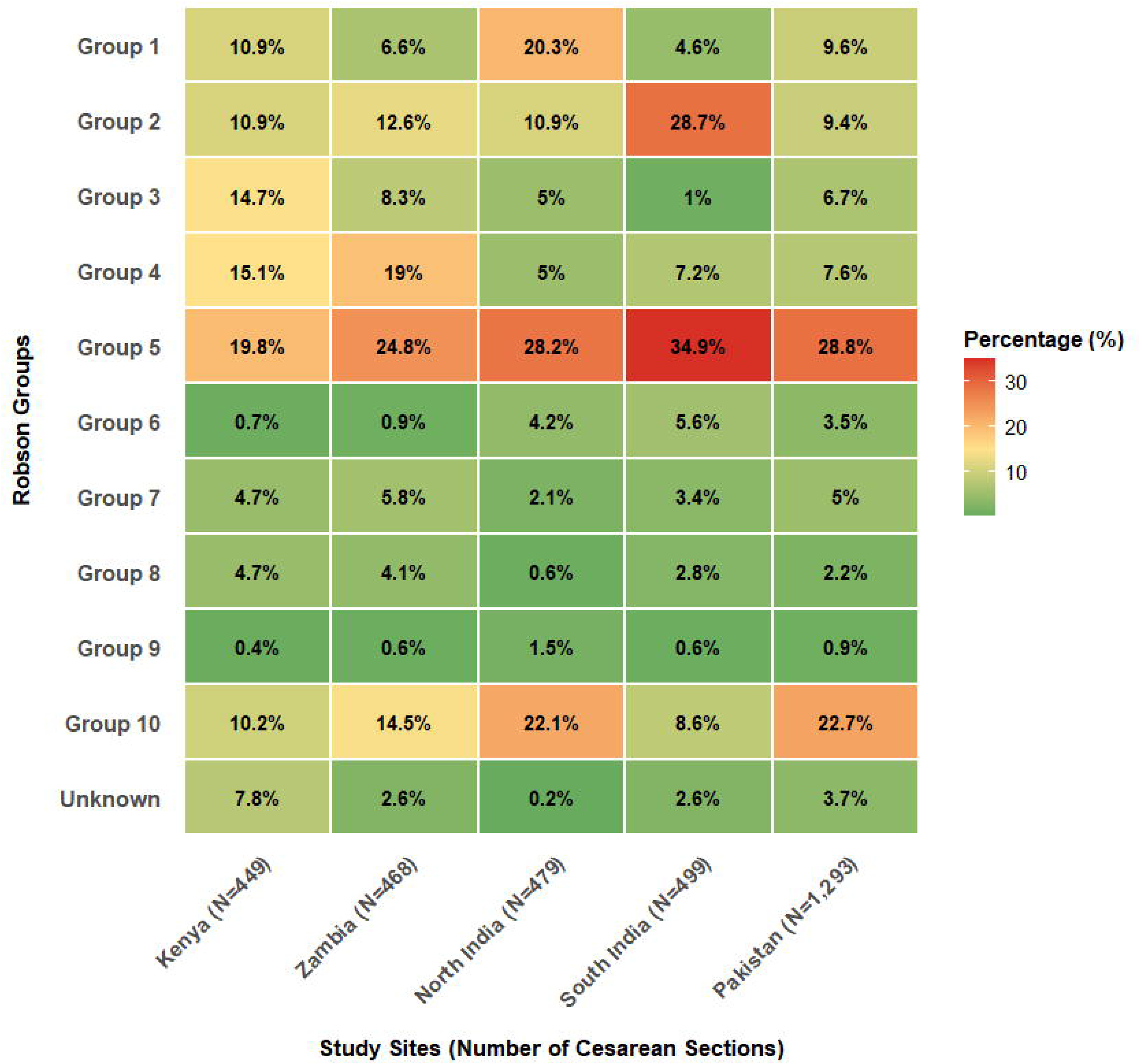
Relative contribution of each Robson Group to the overall proportion of cesarean sections

We found an overall association between CS and risk of adverse maternal and infant outcomes within six weeks postpartum (**Table S5**). Specifically, CS was associated with increased risk of maternal near-miss (RR 2.24 [95% CI 1.90, 2.65]), prolonged hospitalization (RR 2.46 [95% CI 2.23, 2.72]), severe postpartum hemorrhage (RR 2.24 [95% CI 1.99, 2.99]), and NICU admission (RR 2.70 95% CI 2.35, 3.09]) in multivariable adjusted models. There were insufficient neonatal deaths to produce estimates.

CS was associated with at least one adverse maternal outcome within Groups 1, 2, 3, 4, and 10 (**Figure 5, Table S5**). Within Group 1, CS was associated with increased risk of maternal near-miss (RR 1.97 [95% CI 1.16, 3.33]), prolonged hospitalization (RR 4.65 [95% CI 3.55, 6.09]), severe post-partum hemorrhage (RR 1.88 [95% CI 1.01, 3.55]), and NICU admission (RR 2.64 [95% CI 1.78, 3.92]). Within Group 4, CS was associated with increased risk of prolonged hospitalization (RR 2.44 [95% CI 1.81, 3.30]), severe postpartum hemorrhage (RR 2.24 [95% CI 1.11, 4.55]) and NICU admission (RR 2.50 [95% CI 1.54, 4.04]). Within Group 10, CS was associated with increased risk of maternal near-miss (RR 2.64 [95% CI 1.80, 3.86]), prolonged hospitalization (RR 2.47 [95% CI 1.78, 3.42]), severe postpartum hemorrhage (RR 2.67 [95% CI 1.65, 4.31]), and NICU admission (RR 1.47 [95% CI 1.19, 1.84]).

**Figure 5.**
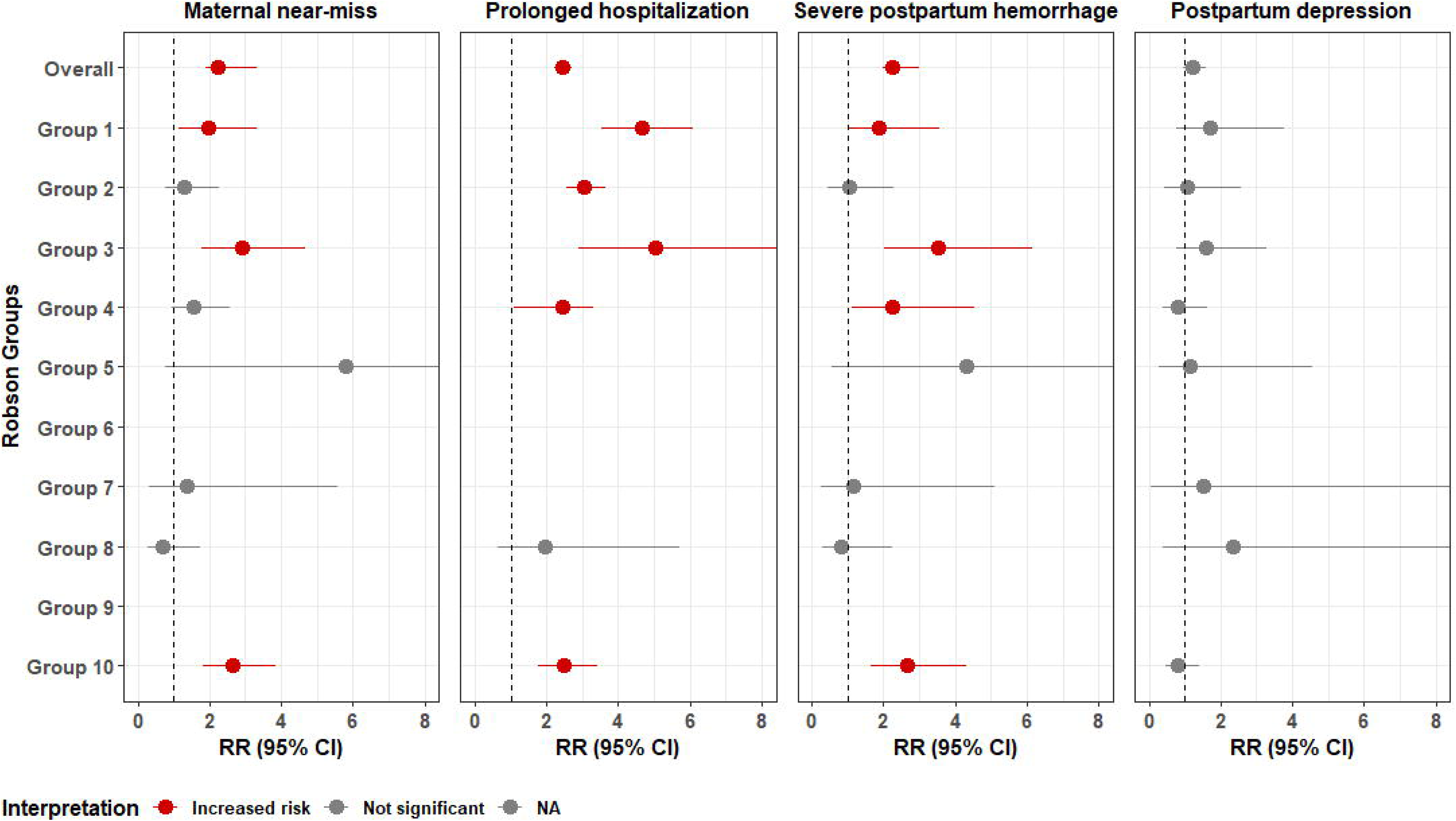
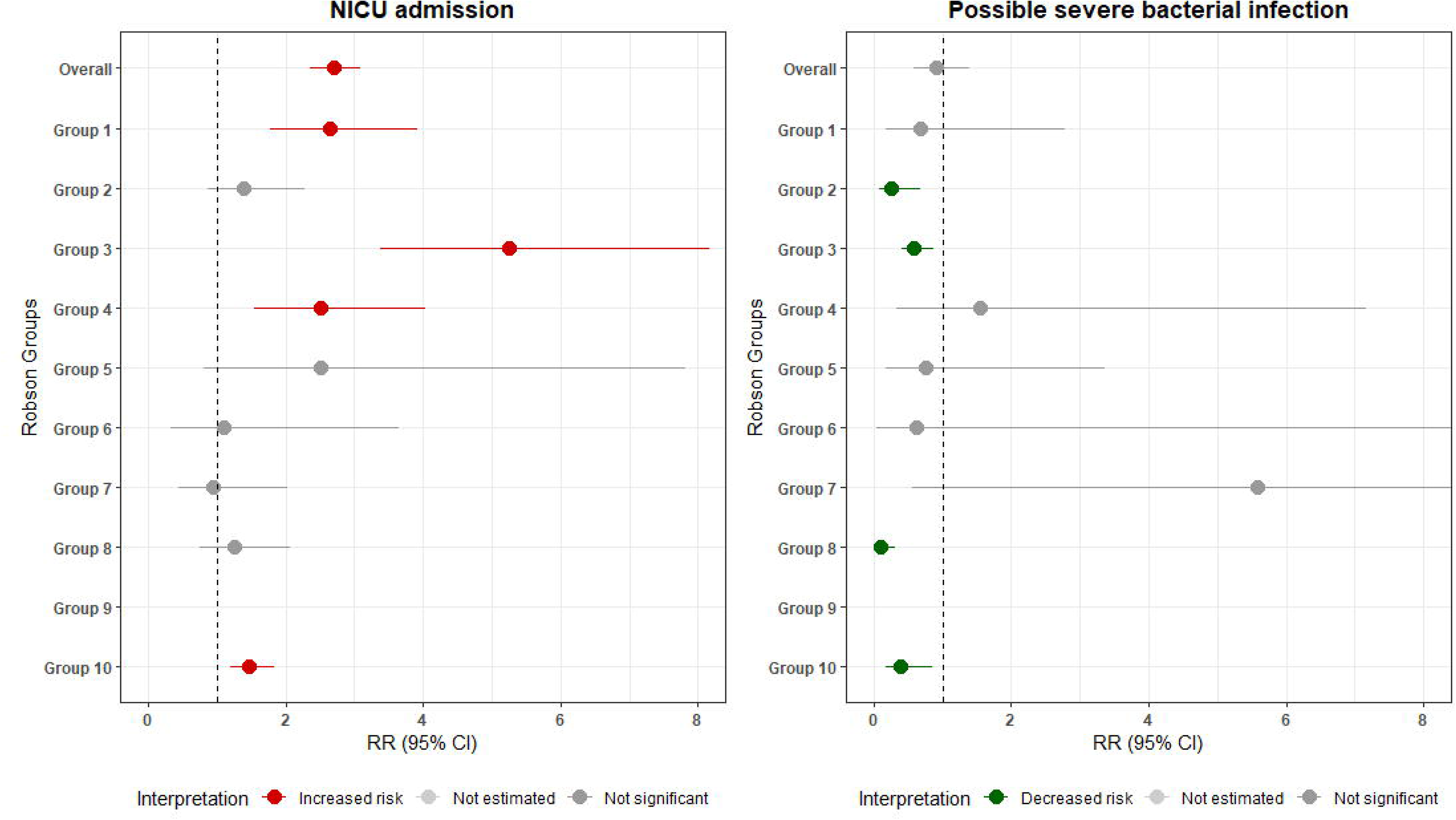
Forest plots of adjusted relative risks (RR) for adverse outcomes associated with cesarean section (ref: vaginal delivery), stratified by Robson Group

Compared to vaginal delivery, CS was associated with lower risk of possible severe bacterial infection in infants within Group 2 (RR 0.26 [95% CI 0.09, 0.68]), Group 3 (RR 0.59 [95% CI 0.41, 0.87]), Group 8 (RR 0.11 [95% CI 0.04, 0.31]) and Group 10 (RR 0.40 [95% CI 0.18, 0.85]). The overall association between CS and possible severe bacterial infection was insignificant.

## DISCUSSION

We examined CS incidence and within-Robson-Group risks of adverse events associated with CS delivery. CS incidences exceeded WHO Robson Group benchmarks, particularly for Group 4 (multipara with induced labor/pre-labor CS), Group 5 (prior CS), and Group 10 (preterm) [11]. There was considerable between-sites variation in group size and group CS incidence within obstetrically similar populations, demonstrating the impact of clinical protocols and facility readiness on CS provision. The magnitude of the association between delivery mode and risk of adverse outcomes varied by Robson Group; broadly, CS-associated risk was greater in lower-risk obstetric groups (Groups 1-4) than higher-risk (Groups 6-9).

We add to an emerging body of evidence linking CS to increased risk of adverse maternal and infant events and found risk was concentrated in Robson Groups 1-4 (singleton, term pregnancies without prior CS) and Group 10 (preterm) [19, 20]. Several explanations warrant consideration. As most CS deliveries in these groups are likely unplanned, the risk may reflect intrapartum complications (i.e. confounding by indication). Alternatively, the CS-associated morbidity in these groups could point to late hospital presentation or delayed service provision, consistent with the Three Delays model and previous research in LMICs [4, 21–23]. Conversely, CS was associated with reduced risk of infant possible severe bacterial infection in multiple groups: plausible as maternal prophylactic antibiotic administration is standard for CS. No association between CS and adverse events was observed for Group 5 (prior CS), possibly demonstrating the safety of planned CS. By comparison, a systematic review estimated that CS was associated with 1.32 (95% CI 1.01, 1.74) higher odds of maternal adverse outcomes in Group 5 [24]. Future studies may compare within-group associations of CS and adverse outcomes by country, as the relationship between CS and hemorrhage or stillbirth may be more pronounced in African versus non-African LMICs [25].

Our analyses revealed substantial between-site variation in both the proportion of participants undergoing labor induction or pre-labor CS and CS incidence following provider-initiated delivery, potentially indicating the influence of facility-specific labor management protocols on operative delivery. Twenty-nine percent of our sample had labor induced: higher than regional induction estimates of 4% in Africa and 12% in Asia, though it may mirror the challenge of generalizing from single-country reviews and the nature of PRISMA study facilities [26, 27]. Although provider-initiated delivery can reduce maternal and fetal risks, labor induction carries risks and evidence is mixed as to whether it increases likelihood of CS [28, 29]. Our results show that induction can be performed successfully, as in the North India site which exhibited the lowest CS incidence among Groups 2 and 4 (nullipara and multipara with induced labor/pre- labor CS). Comparatively, the African sites had two-to-six times greater CS incidence within these groups, possibly reflecting circumstances at lower-resourced facilities that lead to fewer provider-initiated deliveries and higher risk of unsuccessful induction, including inadequate maternal and fetal monitoring and insufficient staff to manage provider-initiated labor [30].

This study benefits from a robust dataset and accurate gestational age dating. To our knowledge, ours is the first publication applying the Robson Classification to facilities in Zambia, and we contribute to a small scholarship from India, Pakistan, and Kenya [31, 32]. As our facilities were not selected randomly, we cannot generalize our findings nationally. Interpretation of results is hindered without information on clinician training and experience level, as well as by the inclusion of facilities that lacked the capacity to carry out CS and those that were referral centers. We were further limited by missing data, with 7% of our sample missing at least one Robson Group criteria and classified as “Unknown.”

## CONCLUSIONS

Our results revealed variation in CS incidence within comparable obstetric populations across four LMICs, most notably for Groups 2 and 4, revealing the importance of facility readiness for labor management in determining delivery mode. We found that CS was associated with increased risk of adverse maternal and newborn outcomes within lower-risk obstetric groups (Robson Groups 1-4) and preterm deliveries (Group 10), but not among higher-risk obstetric groups (Groups 6-9) or prior CS (Group 5). This may be explained by scheduled versus unplanned CS, or late CS due to delays in care-seeking or service provision. Our work emphasizes the value of going beyond overall CS rate to better understand context-specific drivers of CS and areas for quality improvement.

## AUTHOR CONTRIBUTIONS

Conceptualization: SGB, NSY, JA, VA, RT, BJW, NBA, and MBS. Data curation: NSY, JA, PK, KK, BL, EMO, and NS. Formal analysis: NBA and EMO. Funding acquisition: VA, SJB, AGC, ZH, MPK, SM, CNM, WM, MIN, ERS, and MBS. Methodology: SGB, PK, KK, BL, EMO, NS, ERS, NBA, and MBS. Project administration: SGB, NSY, VA, FA, SJB, AGC, ZH, MPK, SM, CNM, WM, MIN, CS, ERS, and MBS. Resources: ERS. Supervision: VA, SJB, AGC, ZH, MPK, SM, CNM, WM, MIN, ERS, and MBS. Visualization: NBA. Writing—original draft: SGB, NSY, JA, VA, RT, NBA, and MBS. Writing—review & editing: SGB, NSY, JA, VA, RT, BJW, FA, AGC, ZH, MPK, PK, SM, WM, KK, BL, EMO, CS, NS, ERS, NBA, and MBS.

## Data Availability

All data produced in the present study are available upon reasonable request to the authors.

## ACKNOWLEDGEMENTS

The authors would like to acknowledge the participants and their families without whom this work would not have been possible. We would also like to thank the research teams at each study site, including: Kephas Otieno, Felix Hayara, Zacchaeus Abaja, Maryanne Nyanjom, Beverly Olang’o, Dorothy Lynda Achieng, Cynthia Ogwang, Dickson Gethi, Fredrick Onduru, Irene Migott, Lydia Ojowi, Janepher Ambuso, Peter Otieno, Vincent Obiero, Mary Omwalo, Harun Owour, and Edwin Kiplagat from the Kenya Medical Research Institute, Kenya; Fyezah Jehan, Amna Khan, Shayan Khakwani, Asad Sheikh, Azqa Mazhar, Iqra Aslam from the Department of Pediatrics and Child Health, The Aga Khan University, Pakistan; Arun Singh Jadaun, Meghna Singh, and Ashwini Kumar from the Society of Applied Studies, New Delhi, India; Lydia, Daniel Jebakumar, and Jasmine Sugirtha from the Christian Medical College of Vellore, Vellore, India; and Inutu Matongo, Rachael Ngulube, and Felistas Mbewe from the University of North Carolina Global Projects Zambia, Lusaka, Zambia. This work was supported by the Gates Foundation. The conclusions and opinions expressed in this work are those of the author(s) alone and shall not be attributed to the Foundation. Under the grant conditions of the Foundation, a Creative Commons Attribution 4.0 License has already been assigned to the Author Accepted Manuscript version that might arise from this submission.

## LIST OF SUPPLEMENTARY TABLES AND FIGURES

Table S1. Definitions of study outcomes and covariates

Table S2. Distribution of birth outcomes and pregnancy complications in the analytic population

Table S3. Robson group size by study sites

Table S4. Cesarean incidences (%) by Robson Group and study site

