## Supplementary Tables for "Moving beyond overall cesarean rates: Evaluating associations of delivery mode and adverse outcomes by Robson Group among the PRISMA Maternal and Newborn Health Study cohort in sub-Saharan Africa and South Asia"

### SUPPLEMENTARY MATERIAL

##### **Table S1. Definitions of study outcomes and covariates**

| **Outcomes** | **Definition** | |
| --- | --- | --- |
| Prolonged hospitalization | >3 days in the hospital after delivery | |
| Severe postpartum hemorrhage | Had ≥1000 mL estimated blood loss or underwent a procedure for hemorrhage | |
| Maternal near-miss | Experienced any of the following after delivery and survived:  • Organ dysfunction  • Hysterectomy  • Postpartum preeclampsia with severe features or eclampsia | • Severe postpartum hemorrhage  • Blood transfusion  • Severe anemia (hemoglobin <7 g/dL)  • Admission to an intensive care unit |
| Postpartum depression | Present if scored ≥11 on the Edinburgh Postnatal Depression Scale at six weeks postpartum | |
| Neonatal death | Mortality from any cause within the first 28 days of life | |
| Possible severe bacterial infection | Presence of any clinical signs or symptoms from World Health Organization Integrated Management of Childhood Illness guidelines within the first six weeks of life [[33]](https://paperpile.com/c/ZNczWy/38HW) | |
| NICU admission | Binary indicator (yes/no) | |
| **Pregnancy Comorbidities** | **Definition** | |
| Anemia | Moderate to severe if hemoglobin <10 g/dL in first or third trimester or <9.5 g/dL in the second trimester [[34]](https://paperpile.com/c/ZNczWy/459R) | |
| Hypertensive disorders of pregnancy | Hypertensive disorders of pregnancy encompassed elevated systolic (≥140 mmHg) and/or diastolic blood pressure (≥90 mmHg) on two separate occasions, one or more high blood pressure readings combined with new receipt of anti-hypertensive medication, or any clinical diagnosis of hypertensive disorder of pregnancy, including chronic hypertension, gestational hypertension, preeclampsia with or without severe features and superimposed pre-eclampsia [[35]](https://paperpile.com/c/ZNczWy/YsPC) | |
| Gestational diabetes mellitus | Fasting glucose ≥5.1 mmol/L, oral glucose tolerance test (OGTT) 1-hour result ≥10.0 mmol/L, or OGTT 2-hour result ≥8.5 mmol/L at approximately 28 weeks gestation, excluding overt diabetes cases [[36]](https://paperpile.com/c/ZNczWy/vKFE). Overt diabetes was defined by HbA1c level ≥6.5% at <20 weeks of gestation, or reported diagnosis of pre-existing diabetes if HbA1c measurement is missing in any visit before ANC 20 weeks. | |

##### **Table S2. Distribution of birth outcomes and pregnancy complications in the analytic population**

| **Characteristic** | **Overall** | **Zambia** | **Kenya** | **North India** | **South India** | **Pakistan** |
| --- | --- | --- | --- | --- | --- | --- |
| Sample size, N | 10,996 | 1,415 | 2,242 | 1,828 | 1,919 | 3,592 |
| Birth outcome* | | | | | | |
| Live birth, % | 99.4 | 98.2 | 99.2 | 99.3 | 99.5 | 100.0 |
| Stillbirth (≥28 weeks), % | 0.6 | 1.8 | 0.8 | 0.7 | 0.5 | 0.0 |
| Diabetes in pregnancy | | | | | | |
| Overt diabetes, % | 1.6 | 4.0 | 0.3 | 0.1 | 2.6 | 1.7 |
| Gestational diabetes, % | 5.7 | 6.6 | 2.3 | 3.9 | 7.9 | 7.0 |
| Hypertensive disorders of pregnancy (chronic hypertension, gestational hypertension, preeclampsia with or without severe features and superimposed preeclampsia) | | | | | | |
| Yes, % | 9.6 | 18.8 | 10.2 | 4.4 | 10.4 | 7.7 |
| No, % | 90.4 | 81.2 | 89.8 | 95.6 | 89.6 | 92.3 |
| Moderate or severe anemia (<10 g/dL in 1st and 3rd trimester and <9.5 g/dL in 2nd trimester ) | | | | | | |
| Yes, % | 45.3 | 38.7 | 59.4 | 51.9 | 12.6 | 53.3 |
| No, % | 54.7 | 61.4 | 40.6 | 48.1 | 87.4 | 46.7 |

**Total number of children = 11,144; Number of children from Zambia=1,445, Kenya=2279, North India=1,839, South India=1,941 and Pakistan=3,640.*

##### **Table S3. Robson group size^1^ by study sites**

| **Robson Group** | **Study Site, n (%)** | | | | |
| --- | --- | --- | --- | --- | --- |
|  | **Kenya** | **Zambia** | **North India** | **South India** | **Pakistan** |
| Group 1 | 438 (19.5) | 179 (12.6) | 335 (18.3) | 277 (14.4) | 495 (13.8) |
| Group 2 | 92 (4.1) | 99 (7.0) | 264 (14.4) | 512 (26.7) | 266 (7.4) |
| Group 3 | 1088 (48.5) | 549 (38.8) | 391 (21.4) | 370 (19.3) | 1,099 (30.6) |
| Group 4 | 199 (8.9) | 204 (14.4) | 372 (20.3) | 376 (19.6) | 449 (12.5) |
| Group 5 | 105 (4.7) | 147 (10.4) | 145 (7.9) | 174 (9.1) | 409 (11.4) |
| Group 6 | 6 (0.3) | 4 (0.3) | 27 (1.5) | 31 (1.6) | 46 (1.3) |
| Group 7 | 35 (1.6) | 31 (2.2) | 18 (1.0) | 19 (1.0) | 82 (2.3) |
| Group 8 | 41 (1.8) | 31 (2.2) | 5 (0.3) | 20 (1.0) | 49 (1.4) |
| Group 9 | 2 (0.1) | 3 (0.2) | 7 (0.4) | 3 (0.2) | 11 (0.3) |
| Group 10 | 146 (6.5) | 137 (9.7) | 263 (14.4) | 116 (6.0) | 637 (17.7) |
| Unknown | 90 (4.0) | 31 (2.2) | 1 (0.1) | 21 (1.1) | 49 (1.3) |
| **Total** | 2,242 (100) | 1415 (100) | 1828 (100) | 1919 (100) | 3592 (100) |

**^1^***Robson group size was calculated by dividing the number of women in the Robson group by the total number of women.*

*^2^Group 1 = Nulliparous women with a single cephalic pregnancy, ≥37 weeks gestation in spontaneous labour; Group 2 = Nulliparous women with a single cephalic pregnancy, ≥37 weeks gestation who had labour induced or were delivered by CS before labour; Group 3 = Multiparous women without a previous CS, with a single cephalic pregnancy, ≥37 weeks gestation in spontaneous labour; Group 4 = Multiparous women without a previous CS, with a single cephalic pregnancy, ≥37 weeks gestation who had labour induced or were delivered by CS before labour; Group 5 = All multiparous women with at least one previous CS, with a single cephalic pregnancy, ≥37 weeks gestation; Group 6 = All nulliparous women with a single breech pregnancy; Group 7 = All multiparous women with a single breech pregnancy including women with previous CS(s); Group 8 = All women with multiple pregnancies including women with previous CS(s); Group 9 = All women with a single pregnancy with a transverse or oblique lie, including women with previous CS(s); Group 10 = All women with a single cephalic pregnancy < 37 weeks gestation, including women with previous CS(s); Unknown= Group status not known due to lack of information on one or more of the required obstetric criterias.*

##### **Table S4. Cesarean incidences (%) by Robson Group and study site^1^**

| **Robson Group^2^** | **Proportion of Group Delivered via Cesarean (n/N, %)** | | | | | |
| --- | --- | --- | --- | --- | --- | --- |
|  | **Overall** | **Kenya** | **Zambia** | **North India** | **South India** | **Pakistan** |
| Group 1 | 325/1724 (18.9%) | 49/438 (11.2%) | 32/179 (17.9%) | 97/335 (29.0%) | 23/277 (8.3%) | 124/495 (25.1%) |
| Group 2 | 425/1233 (34.5%) | 49/92 (53.3%) | 59/99 (59.6%) | 52/264 (19.7%) | 143/512 (27.9%) | 122/266 (45.9%) |
| Group 3 | 220/3497 (6.3%) | 66/1088 (6.1%) | 39/549 (7.1%) | 24/391 (6.1%) | 5/370 (1.4%) | 86/1099 (7.8%) |
| Group 4 | 315/1600 (19.7%) | 68/199 (34.2%) | 89/204 (43.6%) | 24/372 (6.5%) | 36/376 (9.6%) | 98/449 (21.8%) |
| Group 5 | 887/980 (90.5%) | 89/105 (84.8%) | 116/147 (78.9%) | 135/145 (93.1%) | 174/174 (100.0%) | 373/409 (91.2%) |
| Group 6 | 100/114 (87.7%) | 3/6 (50.0%) | 4/4 (100.0%) | 20/27 (74.1%) | 28/31 (90.3%) | 45/46 (97.8%) |
| Group 7 | 140/185 (75.7%) | 21/35 (60.0%) | 27/31 (87.1%) | 10/18 (55.6%) | 17/19 (89.5%) | 65/82 (79.3%) |
| Group 8 | 85/146 (58.2%) | 21/41 (51.2%) | 19/31 (61.3%) | 3/5 (60.0%) | 14/20 (70.0%) | 28/49 (57.1%) |
| Group 9 | 26/26 (100.0%) | 2/2 (100.0%) | 3/3 (100.0%) | 7/7 (100.0%) | 3/3 (100.0%) | 11/11 (100.0%) |
| Group 10 | 556/1299 (42.8%) | 46/146 (31.5%) | 68/137 (49.6%) | 106/263 (40.3%) | 43/116 (37.1%) | 293/637 (46.0%) |
| Unknown | 109/192 (56.8%) | 35/90 (38.9%) | 12/31 (38.7%) | 1/1 (100.0%) | 13/21 (61.9%) | 48/49 (98.0%) |
| **Total** | **3188/10996 (29.0%)** | **449/2242 (20.0%)** | **468/1415 (33.1%)** | **479/1828 (26.2%)** | **499/1919 (26.0%)** | **1293/3592 (36.0%)** |

**^1^***CS rate was calculated by dividing the number of CS in the Robson group by the total number of women in that group.*

*^2^Group 1 = Nulliparous women with a single cephalic pregnancy, ≥37 weeks gestation in spontaneous labour; Group 2 = Nulliparous women with a single cephalic pregnancy, ≥37 weeks gestation who had labour induced or were delivered by CS before labour; Group 3 = Multiparous women without a previous CS, with a single cephalic pregnancy, ≥37 weeks gestation in spontaneous labour; Group 4 = Multiparous women without a previous CS, with a single cephalic pregnancy, ≥37 weeks gestation who had labour induced or were delivered by CS before labour; Group 5 = All multiparous women with at least one previous CS, with a single cephalic pregnancy, ≥37 weeks gestation; Group 6 = All nulliparous women with a single breech pregnancy; Group 7 = All multiparous women with a single breech pregnancy including women with previous CS(s); Group 8 = All women with multiple pregnancies including women with previous CS(s); Group 9 = All women with a single pregnancy with a transverse or oblique lie, including women with previous CS(s); Group 10 = All women with a single cephalic pregnancy < 37 weeks gestation, including women with previous CS(s); Unknown= Group status not known due to lack of information on one or more of the required obstetric criterias.*

##### **Table S5. Association of cesarean, compared to vaginal delivery, with adverse outcomes, stratified by Robson Group**

| Robson Group | Relative risk^1^ (95% CI) for adverse maternal and infant outcomes | | | | | |
| --- | --- | --- | --- | --- | --- | --- |
|  | **Maternal near-miss** | **Prolonged maternal hospitalization** | **Severe postpartum hemorrhage** | **Postpartum depression** | **NICU admission** | **Possible severe bacterial infection** |
| Event Risk | 555/10,446 (5.3%) | 1,003/10,996 (9.2%) | 362/10,988 (3.3%) | 248/9,650 (2.6%) | 801/11,084 (7.2%) | 2,002/11,144 (17.7%) |
| Overall RR | **2.24 (1.90, 2.65)** | **2.46 (2.23, 2.72)** | **2.24 (1.99, 2.99)** | **1.23 (0.95, 1.59)** | **2.70 (2.35, 3.09)** | 0.91 (0.59, 1.39) |
| Stratified by Group | | | | | | |
| Group 1 | **1.97 (1.16, 3.33)** | **4.65 (3.55, 6.09)** | **1.88 (1.01, 3.55)** | 1.71 (0.78, 3.76) | **2.64 (1.78, 3.92)** | 0.69 (0.17, 2.79) |
| Group 2 | 1.29 (0.75, 2.25) | **3.06 (2.56, 3.66)** | 1.03 (0.46, 2.28) | 1.06 (0.44, 2.59) | 1.39 (0.86, 2.28) | **0.26 (0.09, 0.68)** |
| Group 3 | **2.90 (1.79, 4.69)** | **5.04 (2.89, 8.77)** | **3.53 (2.03, 6.15)** | 1.59 (0.77, 3.29) | **5.25 (3.38, 8.16)** | **0.59 (0.41, 0.87)** |
| Group 4 | 1.56 (0.95, 2.56) | **2.44 (1.81, 3.30)** | **2.24 (1.11, 4.55)** | 0.80 (0.39, 1.65) | **2.50 (1.54, 4.04)** | 1.54 (0.33, 7.16) |
| Group 5 | 5.79 (0.77, 43.53) | Not estimated | 4.31 (0.57, 32.59) | 1.13 (0.28, 4.59) | 2.51 (0.81, 7.82) | 0.75 (0.17, 3.37) |
| Group 6 | Not estimated | Not estimated | Not estimated | Not estimated | 1.09 (0.33, 3.64) | 0.62 (0.04, 8.68) |
| Group 7 | 1.35 (0.33, 5.59) | Not estimated | 1.17 (0.27, 5.09) | 1.52 (0.05, 50.03) | 0.94 (0.44, 2.02) | 5.58 (0.87, 35.96) |
| Group 8 | 0.71 (0.29, 1.74) | 1.94 (0.66, 5.70) | 0.82 (0.30, 2.25) | 2.35 (0.38, 14.59) | 1.25 (0.76, 2.07) | **0.11 (0.04, 0.31)** |
| Group 9 | Not estimated | Not estimated | Not estimated | Not estimated | Not estimated | Not estimated |
| Group 10 | **2.64 (1.80, 3.86)** | **2.47 (1.78, 3.42)** | **2.67 (1.65, 4.31)** | 0.81 (0.46, 1.41) | **1.47 (1.19, 1.84)** | **0.40 (0.18, 0.85)** |
| Unknown | 3.46 (0.65, 18.39) | **5.07 (1.39, 18.43)** | 2.49 (0.43, 14.56) | **14.93 (9.69, 23.00)** | **4.02 (1.90, 14.81)** | 1.07 (0.45, 2.50) |

*^1^Relative risks obtained from Generalized Linear Models (GLMs) with log link and Poisson distribution and robust standard error adjusted for maternal age categories (<20 years, 20-34 years, 35+years), mother’s educational attainment (no schooling, 1-4 years of schooling, 5+ years of schooling), body mass index at enrolment (underweight, normal, overweight and obese), mother’s height category (<145 cm, 145 to <150cm, 150 to <155cm and >155cm), wealth quintiles and study sites*
